# Genetic associations between SGLT2 inhibition, DPP4 inhibition or GLP1R agonism and prostate cancer risk: a two-sample Mendelian randomisation study

**DOI:** 10.1101/2024.09.15.24313695

**Authors:** Lin Shen, Yifan Yang, Lei Lu, Oscar Hou In Chou, Quinncy Lee, Tong Liu, Guoliang Li, Shuk Han Cheng, Gary Tse, Jiandong Zhou

## Abstract

**Background:** Epidemiological studies have linked the use of the anti-diabetic medications, sodium-glucose co-transporter-2 inhibitors (SGLT2I), dipeptidyl peptidase-4 inhibitors (DPP4I) and glucagon-like peptide-1 receptor agonists (GLP1RA), with prostate cancer risk. However, these studies cannot infer causality.

**Methods:** This was a two-sample Mendelian randomization (MR) using genome-wide association study data designed to identify causal relationships between SGLT2I, DPP4I or GLP1RA and prostate cancer. Genetic associations with HbA1c and risk of prostate cancer were extracted from IEU Open-GWAS Project database with GWAS id ukb-d-30750_irnt (UK Biobank cohort) and ebi-a-GCST006085 (European Molecular Biology Laboratory’s European Bioinformatics Institute cohort), respectively. The two GWAS datasets chosen were obtained from individuals of European ancestry to minimise potential bias from population stratification. The encoding genes targeted by SGLT2I, DPP4I and GLP1RA were SGC5A2, DPP4 and GLP1R, located in Chr16: 31494323-31502181, Chr2: 162848755-162930904 and Chr6: 39016557-39059079, respectively.

**Results:** A total of 31, 2 and 5 single nucleotide variants (SNVs) were used for SGC5A2, DPP4 and GLP1R. Our MR analysis results supported a causal relationship between genetic variation in SLC5A2 and DPP4 and reduced risk of prostate cancer at the Bonferroni-corrected threshold, with odds ratios (OR) [95% confidence intervals] of 0.47 [0.38-0.58] and 0.35 [0.24-0.53], but not for GLP1R (OR: 1.39 [0.93-2.07]). Sensitivity analyses by the leave-one-out method did not significantly alter the OR for SGLT2I.

**Conclusions:** The two-sample MR analysis found that SGLT2 and DPP4 inhibition, but not GLP1R agonism, was associated with lower risks of developing prostate cancer.

## Introduction

Diabetes mellitus is a common metabolic disorder, predisposing affected individuals to adverse cardiovascular events ^1^, macro- and micro-vascular complications ^2^, and cancer ^3^. Whilst achieving good glycaemic control does not appear to reduce the risk of most cancer types ^4^, different anti-diabetic medications may nevertheless modulate cancer risk. These medications may exert anti-cancer effects that may be independent of their actions on glucose control, acting via anti-inflammatory, anti-oxidative and hormonal or other cellular pathways ^5^. Common second line medications include sodium-glucose co-transporter-2 inhibitors (SGLT2I), dipeptidyl peptidase 4 inhibitors (DPP4I) and glucagon-like peptide-1 receptor agonists (GLP1RA) and their use has been associated with altered risks of overall cancer ^6^, as well as individual subtypes of colorectal ^7^, gastric ^8^, hepatocellular ^9^ and prostate ^10^ cancer. However, such epidemiological studies are susceptible to problems such as confounding and reversed causality. These can be addressed by applying Mendelian randomisation (MR), which uses genetic variants as instrumental variables (IVs) to quantify causal effects between exposures and outcomes.

## Methods

The two-sample MR framework was used to investigate the effects of genetic variants in the SGLT2I, DPP4I and GLP1RA targets on the risk of prostate cancer. Our team has previously used MR to evaluate the causality between different exposures and risks of adverse outcomes ^11,12^. Due to the unavailability of GWAS data on the exposure (protein targets of antidiabetic drugs) and considering that lowering of glycated haemoglobin (HbA1c) level is a physiological response to antidiabetic drug, HbA1c was used. Genetic associations with HbA1c and risk of prostate cancer were extracted from IEU Open-GWAS Project database with GWAS id ukb-d-30750_irnt (UK Biobank cohort) and ebi-a-GCST006085 (European Molecular Biology Laboratory’s European Bioinformatics Institute cohort), respectively. The two GWAS datasets chosen were obtained from individuals of European ancestry to minimise potential bias from population stratification.

The IVs were screened for the genes that encode antidiabetic drug target proteins. Variations within and around these genes were used to assess the effects of drug use on exposure and outcomes. The detailed information about the protein targets and encoding genes for SGLT2I, DPP4I, and GLP1RA is provided in **Table 1**. SNVs within each encoding gene (within ±100kb base pairs of the gene location) were identified. Variants with p-values greater than 5*10-4 and a minor allele frequency (MAF) below 0.01 in the GWAS data for HbA1c were excluded. The remained variants for each antidiabetic drug target were selected as IVs and clumped with an R2 of 0.8, a window size of 250 kb, and the reference population set to EUR.

**Table 1.** Detailed information of the different anti-diabetic drug classes, targets, and encoding genes.

| Drug class | Drug target<br>(*Encoding gene*) | Gene region |
| --- | --- | --- |
| Sodium-glucose cotransporter 2 (SGLT2) inhibitors | SGLT2 (*SGC5A2*) | Chr16: 31494323-31502181 |
| Dipeptidyl peptidase 4 (DPP-IV) inhibitors | DPP-IV (*DPP4*) | Chr2: 162848755-162930904 |
| Glucagon-likepeptide 1 (GLP1) agonists | GLP-1R (*GLP1R*) | Chr6: 39016557-39059079 |

To ensure the validity of the MR analyses, three conditions must be satisfied ^13^. 1) Relevance: the IVs are associated with the risk factor of interest. It was achieved by selecting IVs with a F-statistic >10, 2) Independence, the IVs do not share common cause with the outcome. This was fulfilled by random assortment of genetic variants during conception. 3) Exclusion restriction, where IVs do not affect outcomes except through the risk factor. This assumption was tested by Cochran Q test, exclusion restriction, which evaluates heterogeneity. Evaluation of horizontal pleiotropy was conducted using the MR Egger method.

In instances where all three assumptions were met, the Inverse Variance Weighted (IVW) method was applied to yield unbiased estimates on the associations between exposures and outcomes. The F-statistic is calculated by dividing the square of beta by the square of the standard error, where an F-statistic >10 indicated sufficient instrument strength. Following Bonferroni correction, the significance level of multiple testing for 3 drugs classes was set as P< 0.017 (0.05/3).

The leave-one-out method was selected for sensitivity analysis, which entailed systematically excluding each variant within the drug’s target gene region and subsequently calculating the meta-effects of the remaining variants. The absence of statistically significant changes in MR estimates following the removal of each variant implies the robustness of the obtained results. All analyses were performed in R (Version 4.3.1), using the TwoSampleMR (Version: 0.5.7) package.

## Results

For the primary analysis, 31 variants were selected for SGLT2 inhibition (mean F-statistic of 27), 2 for DPP-4 inhibition (mean F-statistic of 17), and 25 for GLP-1 agonism (mean F-statistic of 20). Detailed data of the IVs to proxy antidiabetic drugs are shown in **Table 2**. The results of the MR analyses are shown in **Figure 1**. SGLT2I, as reflected by genetic variations in SLC5A2, led to reduced risks of prostate cancer at the Bonferroni-corrected threshold (odds ratio [OR] = 0.47, 95% CI 0.38-0.58). DPP4 inhibition also led to reduced risks of prostate cancer (OR = 0.35, 95% CI 0.24-0.53). However, no significant protective effect was observed for GLP1R agonism (OR: 1.39 [0.93-2.07]). No heterogeneity or pleiotropy was detected within the IVs in SLC5A2 and DPP4 analysis **(Figure 1)**. More details on the MR analysis can be found in **Table 3**. Sensitivity analysis using the leave-one-out was performed for the genetic variations in SLC5A2 and the risk of prostate cancer. The analyses demonstrated the robustness of the results, indicating that after excluding each SNV, the overall error lines showed minimal changes **(Figure 2)**. However, due to the insufficient genetic variants identified for DPP4, sensitivity analysis was not performed.

**Figure 1.**
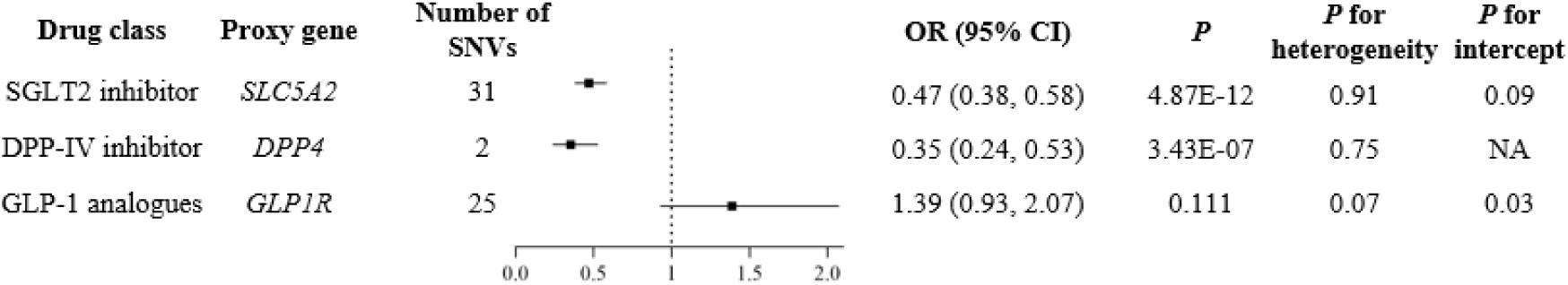
Estimated Effects of Genetic Variation in Antidiabetic Drug Targets on the risk of prostate cancer. Proxy gene is the gene that encodes the drug target proteins. *P* < 0.01 indicates statistical significance, and 0.01 < *p* < 0.05 indicates suggestive significance. *P* for heterogeneity < 0.05 indicates possible heterogeneity, whereas *p* for intercept <0.05 indicates substantial bias from pleiotropy. IVs, instrumental variables; SGLT2I, sodium-glucose cotransporter 2; DPP4I, dipeptidyl peptidase 4; GLP1a, glucagon-likepeptide 1; SNVs, single-nucleotide variations.

**Figure 2.**
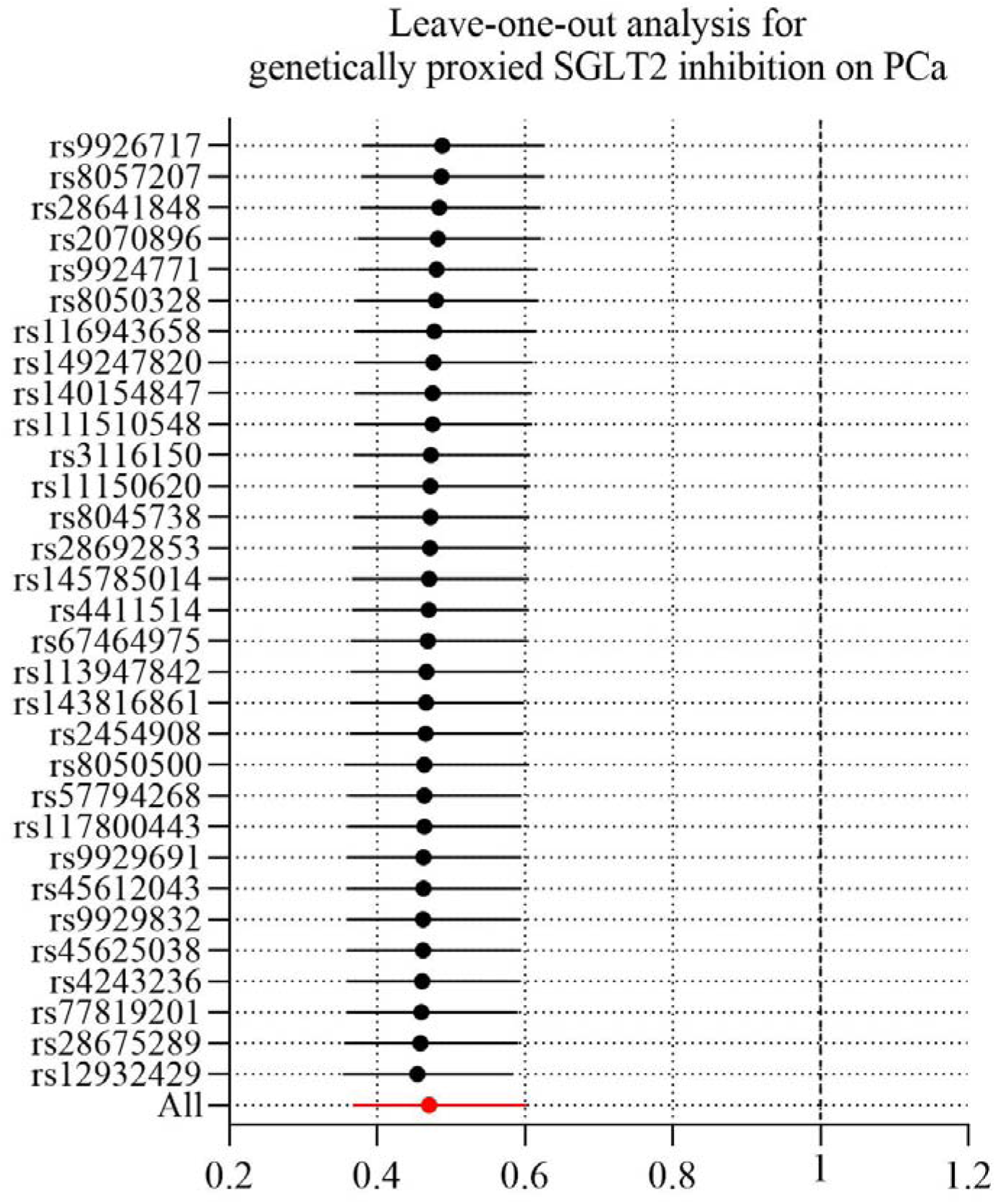
Leave-one-out sensitivity analysis for genetically proxied SGLT2 inhibition on PCa. SGLT2, sodium-glucose cotransporter 2; PCa, Prostate cancer.

**Table 2.** Genetic variants used as instrumental variables for different antidiabetic drug targets for the primary analysis. EA/OA, effect allele/other allele; MAF, minor allele frequency.

| Target | SNP | EA/OA | MAF | Beta | SE | P-value | F |
| --- | --- | --- | --- | --- | --- | --- | --- |
| SGLT2 | rs140154847 | G/T | 0.0151 | 0.0457 | 0.0099 | 3.66E-06 | 21 |
| SGLT2 | rs8050500 | C/T | 0.4460 | -0.0266 | 0.0023 | 1.15E-30 | 121 |
| SGLT2 | rs117800443 | A/G | 0.0696 | 0.0221 | 0.0047 | 2.21E-06 | 22 |
| SGLT2 | rs45625038 | T/C | 0.0296 | 0.0413 | 0.0068 | 1.22E-09 | 34 |
| SGLT2 | rs4243236 | C/G | 0.0540 | -0.0237 | 0.0051 | 3.84E-06 | 20 |
| SGLT2 | rs28692853 | A/C | 0.4926 | -0.0145 | 0.0023 | 2.78E-10 | 36 |
| SGLT2 | rs116943658 | A/G | 0.3299 | -0.0126 | 0.0025 | 3.01E-07 | 24 |
| SGLT2 | rs143816861 | T/C | 0.0181 | -0.0412 | 0.0094 | 1.09E-05 | 21 |
| SGLT2 | rs9926717 | G/A | 0.2844 | -0.0113 | 0.0026 | 9.61E-06 | 18 |
| SGLT2 | rs57794268 | T/C | 0.0663 | 0.0181 | 0.0046 | 8.76E-05 | 14 |
| SGLT2 | rs4411514 | A/G | 0.4429 | -0.0112 | 0.0023 | 1.23E-06 | 21 |
| SGLT2 | rs2070896 | C/T | 0.3750 | -0.0173 | 0.0024 | 1.7E-12 | 48 |
| SGLT2 | rs77819201 | G/A | 0.0270 | 0.0310 | 0.0073 | 2.27E-05 | 17 |
| SGLT2 | rs9929832 | T/C | 0.4738 | -0.0091 | 0.0023 | 9.33E-05 | 14 |
| SGLT2 | rs8050328 | G/T | 0.3637 | -0.0163 | 0.0024 | 1.1E-11 | 42 |
| SGLT2 | rs111510548 | C/T | 0.1034 | -0.0152 | 0.0038 | 6.69E-05 | 15 |
| SGLT2 | rs45612043 | C/A | 0.0445 | -0.0353 | 0.0056 | 2.51E-10 | 36 |
| SGLT2 | rs113947842 | T/G | 0.4066 | -0.0110 | 0.0025 | 1.17E-05 | 20 |
| SGLT2 | rs28641848 | T/C | 0.2793 | -0.0112 | 0.0026 | 1.21E-05 | 18 |
| SGLT2 | rs12932429 | C/T | 0.0497 | -0.0252 | 0.0054 | 2.74E-06 | 21 |
| SGLT2 | rs149247820 | T/C | 0.0151 | 0.0433 | 0.0097 | 7.89E-06 | 19 |
| SGLT2 | rs145785014 | T/C | 0.0534 | -0.0276 | 0.0052 | 9.23E-08 | 26 |
| SGLT2 | rs9924771 | G/A | 0.3482 | -0.0116 | 0.0025 | 4.02E-06 | 21 |
| SGLT2 | rs11150620 | C/G | 0.3015 | -0.0130 | 0.0025 | 2.9E-07 | 25 |
| SGLT2 | rs2454908 | T/C | 0.4354 | -0.0098 | 0.0023 | 2.56E-05 | 16 |
| SGLT2 | rs8045738 | G/T | 0.3065 | -0.0104 | 0.0025 | 2.94E-05 | 16 |
| SGLT2 | rs28675289 | T/C | 0.0444 | -0.0382 | 0.0057 | 1.45E-11 | 43 |
| SGLT2 | rs3116150 | A/G | 0.2411 | 0.0136 | 0.0027 | 4.16E-07 | 23 |
| SGLT2 | rs8057207 | T/C | 0.3622 | -0.0131 | 0.0024 | 4.55E-08 | 27 |
| SGLT2 | rs67464975 | T/C | 0.4699 | -0.0117 | 0.0023 | 4.03E-07 | 24 |
| SGLT2 | rs9929691 | T/C | 0.1501 | 0.0155 | 0.0032 | 1.45E-06 | 21 |
| DPP4 | rs561174039 | A/C | 0.0205 | -0.0364 | 0.0084 | 1.42E-05 | 18 |
| DPP4 | rs78818053 | A/G | 0.0306 | -0.0287 | 0.0067 | 1.62E-05 | 17 |
| GLP1R | rs61086156 | G/C | 0.0156 | 0.0372 | 0.0093 | 6.45E-05 | 15 |
| GLP1R | rs10305420 | T/C | 0.3935 | -0.0111 | 0.0024 | 2.96E-06 | 20 |
| GLP1R | rs9462476 | G/A | 0.3246 | -0.0137 | 0.0025 | 2.69E-08 | 28 |
| GLP1R | rs1544935 | G/T | 0.2162 | -0.0122 | 0.0028 | 1.37E-05 | 17 |
| GLP1R | rs10305423 | T/C | 0.0337 | -0.0286 | 0.0066 | 1.37E-05 | 18 |
| GLP1R | rs1854961 | A/C | 0.4094 | 0.0097 | 0.0023 | 3.04E-05 | 16 |
| GLP1R | rs10305518 | G/T | 0.0568 | 0.0289 | 0.0050 | 7.16E-09 | 31 |
| GLP1R | rs57143316 | C/T | 0.0480 | 0.0294 | 0.0054 | 5.27E-08 | 27 |
| GLP1R | rs10947785 | A/G | 0.4579 | -0.0091 | 0.0023 | 8.04E-05 | 14 |
| GLP1R | rs73731704 | A/G | 0.0581 | 0.0211 | 0.0049 | 1.8E-05 | 17 |
| GLP1R | rs10305442 | G/A | 0.4671 | 0.0095 | 0.0023 | 4.23E-05 | 15 |
| GLP1R | rs6929303 | T/C | 0.0536 | 0.0202 | 0.0051 | 7.66E-05 | 14 |
| GLP1R | rs62688960 | C/T | 0.0322 | -0.0265 | 0.0068 | 9.1E-05 | 15 |
| GLP1R | rs12528717 | T/A | 0.0926 | 0.0186 | 0.0040 | 2.61E-06 | 20 |
| GLP1R | rs10305457 | T/C | 0.0938 | 0.0218 | 0.0039 | 3.53E-08 | 28 |
| GLP1R | rs6458097 | T/C | 0.4437 | -0.0096 | 0.0023 | 3.58E-05 | 16 |
| GLP1R | rs228824 | C/T | 0.3315 | 0.0114 | 0.0025 | 3.81E-06 | 20 |
| GLP1R | rs56384501 | A/G | 0.3520 | 0.0104 | 0.0024 | 1.45E-05 | 17 |
| GLP1R | rs56298410 | T/C | 0.0393 | 0.0275 | 0.0060 | 4.08E-06 | 20 |
| GLP1R | rs9470964 | G/A | 0.0497 | 0.0277 | 0.0054 | 2.27E-07 | 25 |
| GLP1R | rs112901579 | T/C | 0.0315 | 0.0275 | 0.0066 | 3.24E-05 | 16 |
| GLP1R | rs77955327 | C/A | 0.0438 | 0.0285 | 0.0056 | 4.19E-07 | 23 |
| GLP1R | rs9296291 | C/T | 0.2278 | -0.0138 | 0.0028 | 5.27E-07 | 23 |
| GLP1R | rs200781234 | G/A | 0.1074 | 0.0170 | 0.0037 | 5.09E-06 | 19 |
| GLP1R | rs113120569 | A/G | 0.1686 | 0.0177 | 0.0031 | 9.53E-09 | 30 |

**Table 3.**
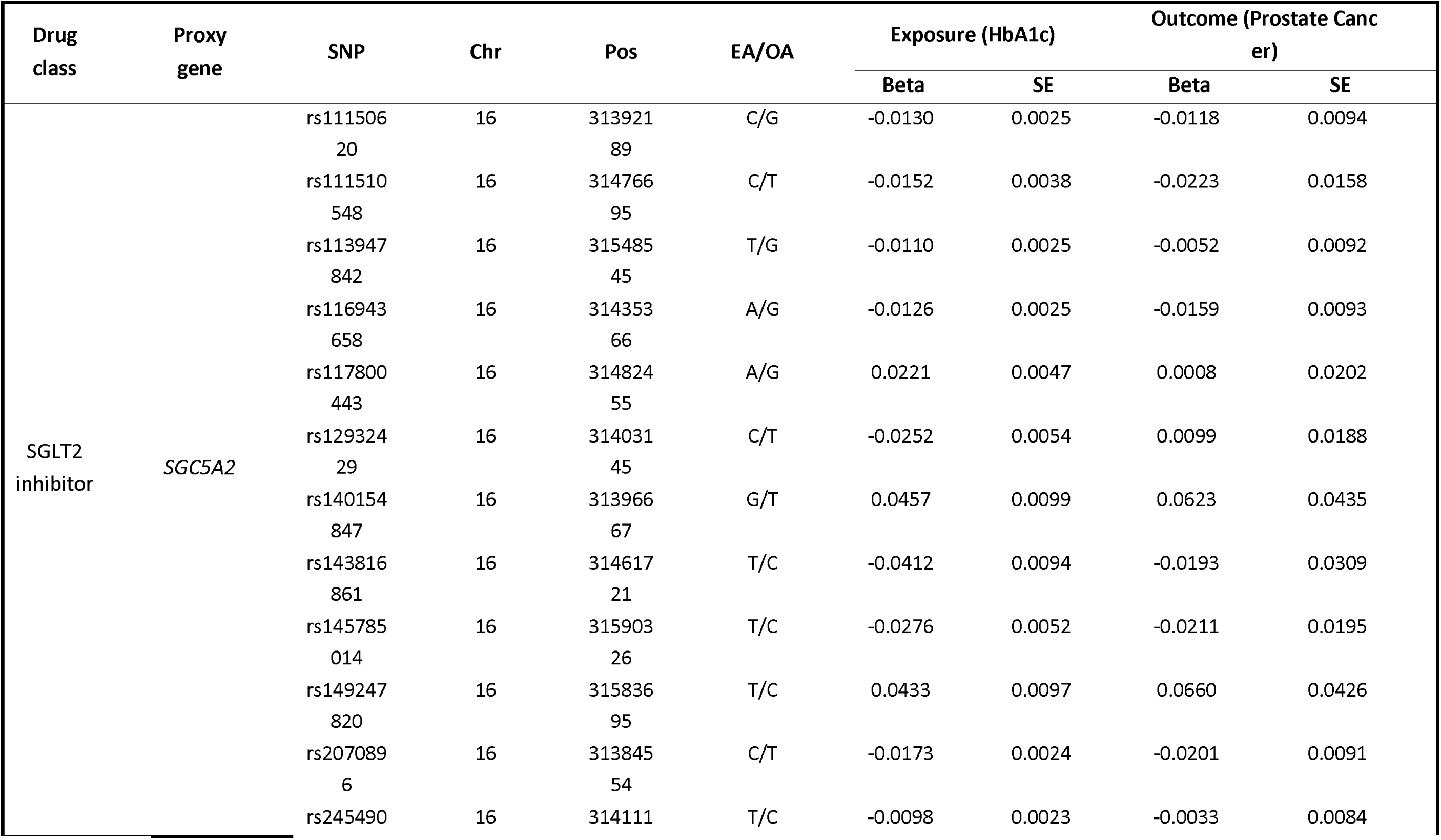
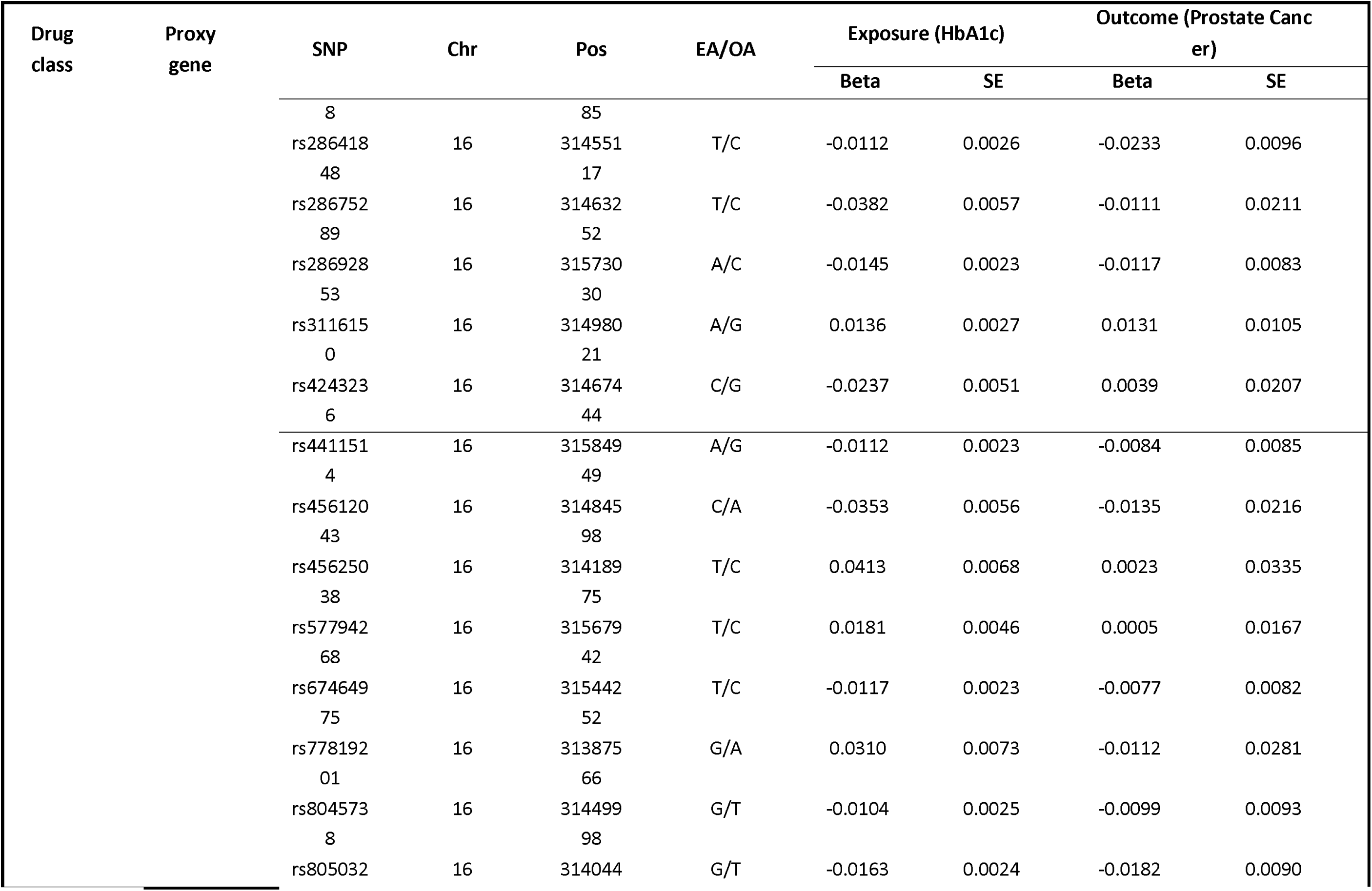
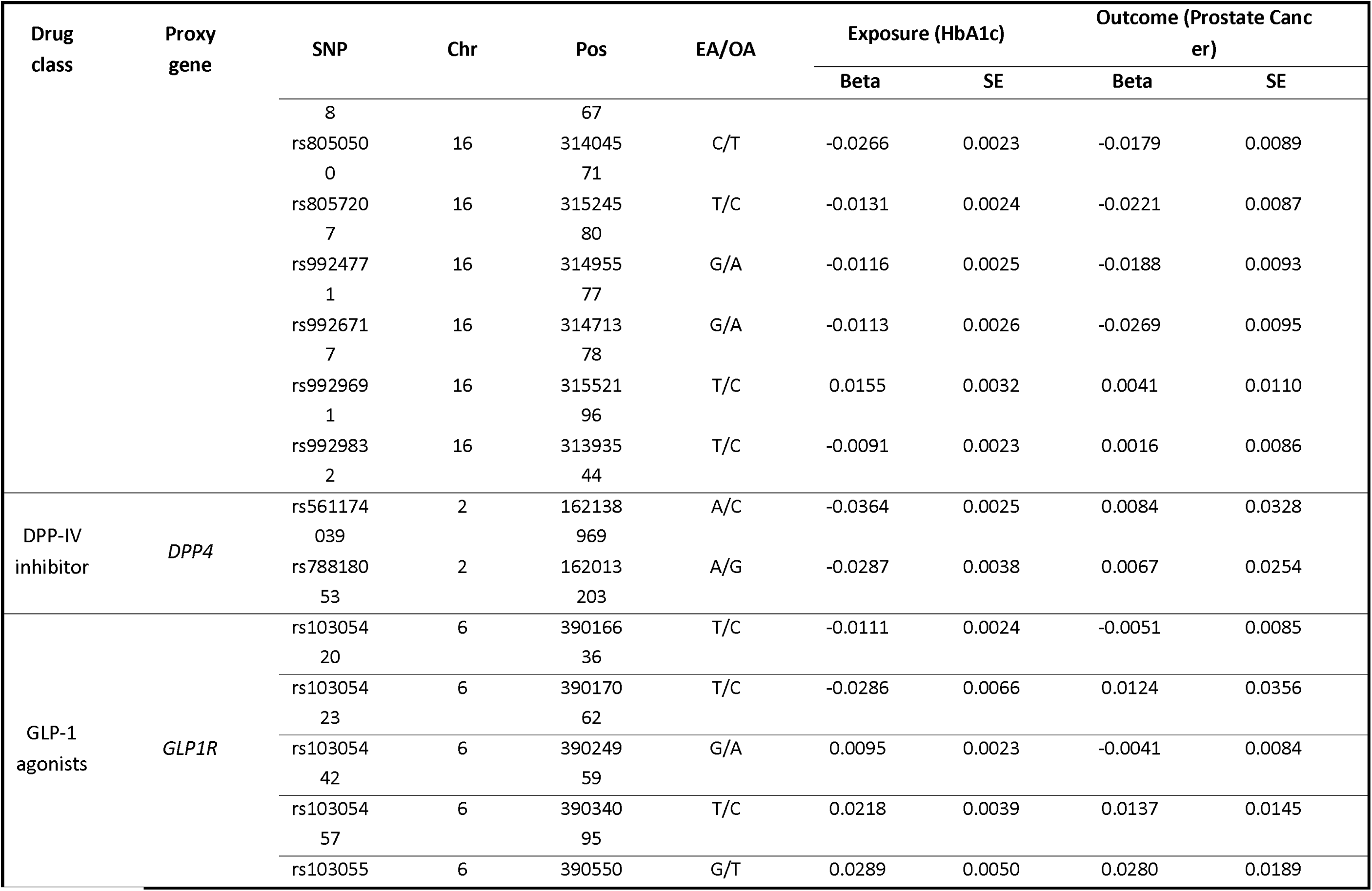
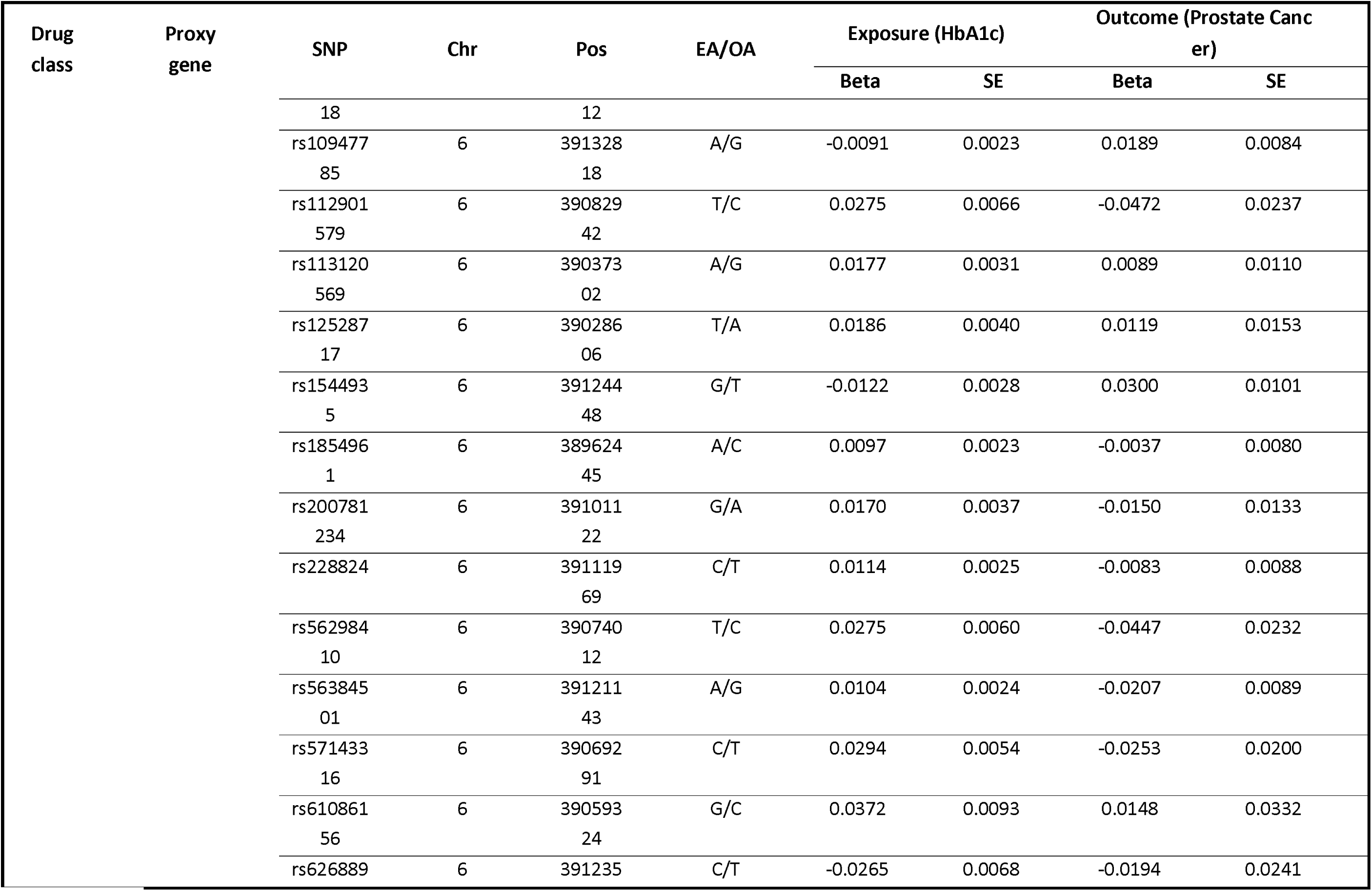
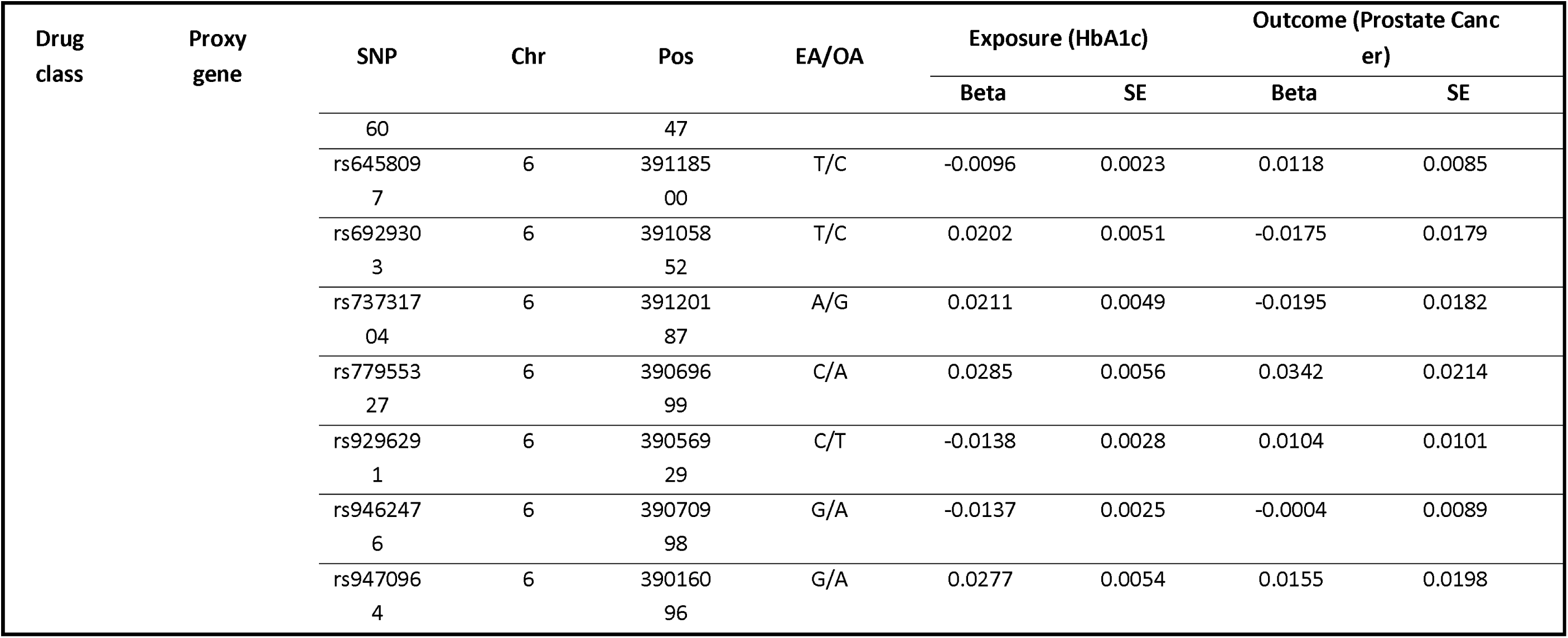
Characteristics of instrumental variables for each drug target in the HbA1c and prostate cancer datasets. Chr, chromosome; EA/OA, effect allele/other allele; Pos, position.

## Discussion

The present two-sample MR study found a significant association between genetic variation in SLC5A2 and DPP4 with prostate cancer. Thus, this study provides evidence that SGLT2 and DPP4 inhibition is protective against prostate cancer. They may directly on the prostate with glycaemic-independent effects. Such findings support possible drug repurposing of SGLT2I and DPP4I on prostate cancer by making use of their pleiotropic effects ^14–16^. Interests in this area was prompted largely due to the fact that cancer cells utilise glucose rather than fatty acids as a primary source of fuel ^17^. Moreover, SGLT2I usage may inhibit the AMPK/mTOR pathway and induce apoptosis of cancerous cells ^18^. SGLT2 inhibition has been shown to reduce glucose uptake and tumour progression in a xenograft model ^19^. SGLT2I inhibited the clonogenic survival of prostate cancer cells ^20^. In our study, the protective effects of SGLT2I were significant amongst patients without comorbidities but not those with prior comorbidities (heart failure, prior acute myocardial infarction, atrial fibrillation, peripheral vascular diseases). Whilst this could be explained by the limited sample size in the subgroups, we may also hypothesise that this could be explained partially by the SGLT2 expression level, as systemic diseases were suggested to increase the expression and activity of SGLT2 ^21^.

Our findings from MR on the protective effects of SGLT2 and DPP4 inhibition on the risks of developing prostate cancer amongst T2DM patients agree with those from other MR studies or observational studies. Regarding SGLT2I, a recent study by our team found that SGLTI use was associated with 55% lowered risk of prostate cancer development compared to DPP4I ^10^. The effect estimate is similar in magnitude to the OR of 0.47 calculated from the MR analyses. For DPP4I, saxagliptin is linked to lower prostate cancer risk compared to sulfonylurea use ^22^. In another observational study, sitagliptin was associated with lower risks of prostate cancer amongst Asians with an odd ratio of 0.61 ^23^. Such observations are in line with OR of 0.35 for DPP4I. Regarding GLP1R agonists, previous MR analyses found neutral effects on different cancer types including ovarian, lung and thyroid cancers ^24^. Liraglutide Effect and Action in Diabetes: Evaluation of Cardiovascular Outcome Results (LEADER) trial revealed that malignant prostate neoplasms were present in a lower proportion of diabetic patients who use liraglutide compared to the placebo group ^25^. In a a nationwide register-based cohort study from Denmark, GLP1RA use was inversely associated with prostate cancer risk compared to basal insulin use ^26^.

### Limitations

Several limitations of this study should be noted. Firstly, the MR analysis was conducted using European populations, which the genetic association might be different from the Hong Kong population in the drug cohort. Lastly, the retrospective design of our study necessitates the presentation of associations but not causal links between SGLT2I versus DPP4I use and the risk of new-onset prostate cancer. Therefore, further research is warranted to explore the beneficial effects of SGLT2I.

## Conclusions

The two-sample MR analysis found that SGLT2 and DPP4 inhibition, but not GLP1R agonism, was associated with lower risks of developing prostate cancer.

## Data Availability

All data produced in the present work are contained in the manuscript

---

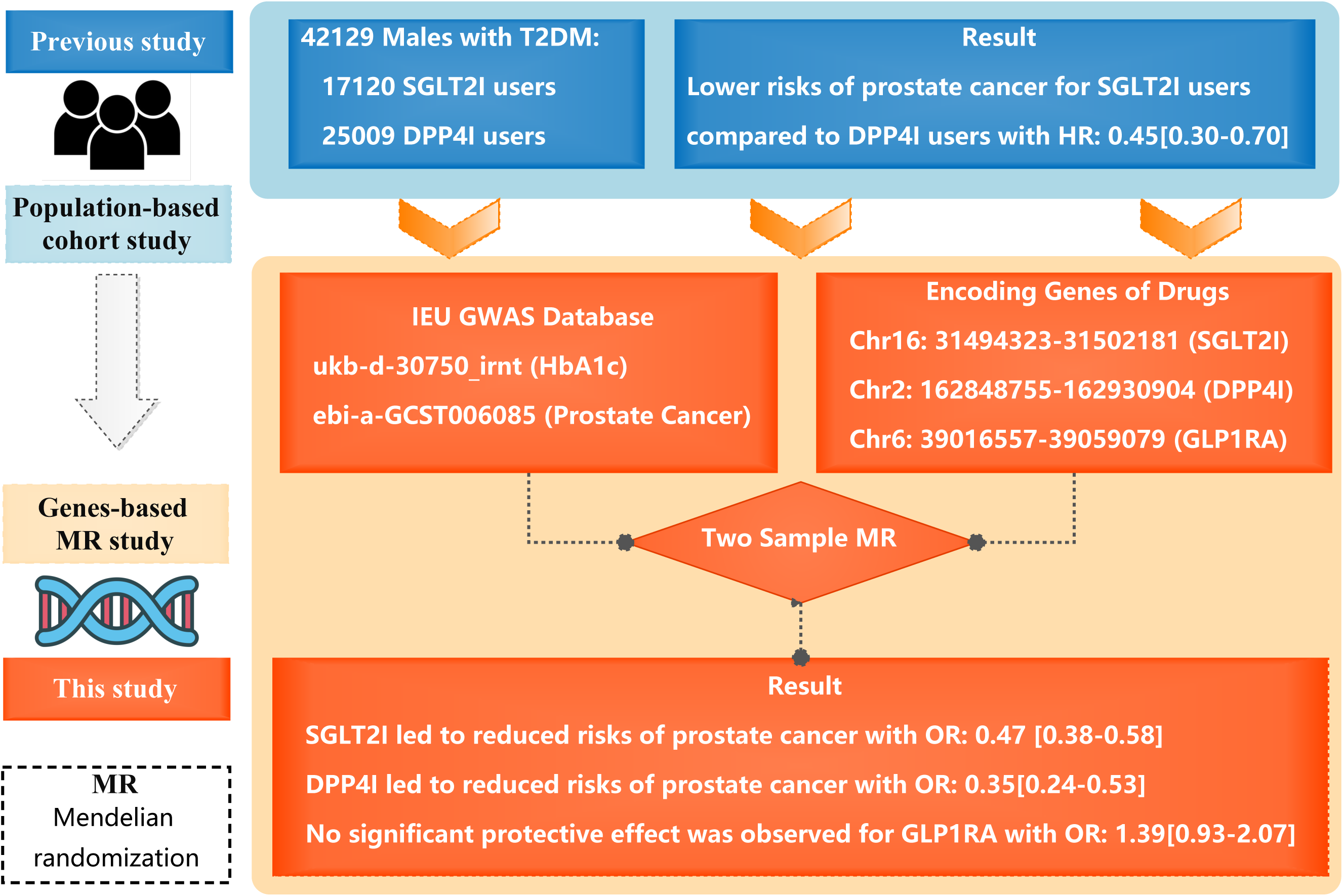

